# Antibody Tests: They Are More Important Than We Thought

**DOI:** 10.1101/2021.02.06.21251251

**Authors:** Luís Guimarães

**Affiliations:** Queen’s University Belfast

**Keywords:** COVID-19, Antibody Tests, Health State Uncertainty, Non-Pharmaceutical Interventions, D62, E17, I12, I18

## Abstract

Antibody testing is a non-pharmaceutical intervention – not recognized so far in the literature – to prevent COVID-19 contagion. I show this in a simple economic model of an epidemic in which agents choose social activity under health state uncertainty. In the model, susceptible and asymptomatic agents are more socially active when they *think* they might be immune. And this increased activity escalates infections, deaths, and welfare losses. Antibody testing, however, prevents this escalation by revealing that those agents are not immune. Through this mechanism, I find that antibody testing prevents about 12% of COVID-19 related deaths within 12 months.

## 1 Introduction

The COVID-19 pandemic has virtually stopped the world economy and has led to the death of more than one million people worldwide by October 2020. In the hope of constraining the pandemic, governments restricted movement, imposed lockdowns and quarantines, forced the closure of businesses, and increased the scale of viral testing and contact-tracing. In this paper, I show that there is another non-pharmaceutical intervention (NPI) to lower contagion: large-scale antibody testing.

There are two obvious reasons to support antibody tests. First, antibody surveys help in understanding the extent of the pandemic, its infection-fatality rate, the duration of immunity, and the proportion of asymptomatic. They have been conducted in several countries and have guided policy and the calibration of epidemiological models. Second, by identifying immune individuals, large-scale antibody testing may facilitate reopening the economy after lockdown (e.g., by issuing the so-called “immunity passports”; see Alvarez, Argente and Lippi, 2020). My paper identifies a new – not as evident – reason to support large-scale antibody testing. By revealing that susceptible and asymptomatic individuals are not immune, antibody testing reduces their social activity lowering the scale of the pandemic.

In the context of the COVID-19 pandemic, most individuals act without knowing their health state. More than half of the infected are asymptomatic (see references in Eikenberry et al., 2020) and, among the symptomatic, many only develop mild symptoms. Furthermore, more than 80% of immune individuals in France, Spain, and the UK are not identified.^1^ Thus, many individuals act without knowing whether they are susceptible, asymptomatic, or immune. Antibody tests end part of this uncertainty by revealing whether individuals are immune or not.^2^

In this paper, I build a simple economic model in which agents maximize their lifetime utility under health state uncertainty. In the model, following Farboodi, Jarosch and Shimer (2020), Garibaldi, Moen and Pissarides (2020), Keppo et al. (2020), and Toxvaerd (2020), agents directly choose social activity, which is an overarching concept covering social and economic returns.^3^ Furthermore, agents can be susceptible, asymptomatic, symptomatic, recovered, or dead. Susceptible agents do not have antibodies against the virus, which puts them at risk of an infection. Asymptomatic and symptomatic agents are infected and may infect others but only symptomatic agents are aware of the infection; thus, asymptomatic agents behave as susceptible agents. Recovered agents have antibodies against COVID-19 and are, thus, immune. Individuals’ optimal social activity depends on their health state and uncertainty. In a world with perfect information, susceptible agents would constrain social activity to reduce exposure to the virus, while recovered agents would not. But, under health state uncertainty, agents rely on expectations of their health state to choose social activity: recovered agents may restrain themselves excessively while, crucial for contagion, susceptible and asymptomatic agents may be excessively active.

Numerical simulations of my model suggest that large-scale antibody testing substantially raises welfare and saves lives. In the model, optimal behavior critically depends on access to antibody tests. As the probability of being immune builds up, susceptible and asymptomatic agents without access to antibody tests become more active than those with access because the former agents think they might be immune whereas the latter know that they are not. Therefore, increasing the scale of antibody testing lowers social activity and contagion by raising the share of agents that are sure of not being immune. Through this channel and using my preferred calibration, antibody testing avoids about 12% of COVID-19 related deaths in 12 months.

Further numerical simulations of my model also suggest that part of the benefits of antibody testing are due to postponing infections to gain time for a vaccine or cure to arrive. Yet, even if a vaccine or cure never arrives, antibody testing permanently lowers contagion; in the long run, antibody testing prevents about 3.4% of COVID-19 related deaths. Furthermore, my simulations also suggest that large-scale antibody testing still substantially lowers contagion and prevents COVID-19 related deaths even if the tests are imperfect.

### Related Literature

The channel through which antibody tests affect behavior in my model has support in data. Gong (2015) assesses how HIV testing affects behavior in Sub-Saharan Africa and documents that individuals surprised by a negative HIV test decrease their risky sexual exposure. Similarly, in my model, agents who think that they are immune and test negative for antibodies against COVID-19 restrain their social activity.

The effects of viral testing in the context of COVID-19 have attracted the attention of the literature (Berger, Herkenhoff and Mongey, 2020; Brotherhood et al., 2020; Eichenbaum, Rebelo and Trabandt, 2020*b*; Piguillem and Shi, 2020).^4^ These papers, and in particular Eichenbaum, Rebelo and Trabandt (2020*b*), offer an enlightening lesson: viral testing only increases welfare if infected agents are quarantined because they no longer face exposure risk. Viral testing, however, differs from antibody testing. A viral test, i.e., a test identifying the current presence of an infection, allows individuals to distinguish the states of susceptible and infected without informing individuals of a past infection. An antibody test, i.e., a test identifying the past presence of an infection, allows individuals to know whether they are immune and to distinguish the recovered state from all the other states without informing individuals of a current infection. This distinction has critical implications because, in contrast with viral testing, I find that antibody testing always increases welfare. This is because positive antibody tests allow recovered agents to enjoy more social activity and negative antibody tests reduce the social activity of those that matter in the propagation of the virus – susceptible and infected agents.

Finally, my paper relates to the long economics literature emphasizing how incentives and individual decision-making shape the extent and welfare costs of an epidemic. In the context of the AIDS pandemic, some examples are Philipson and Posner (1993), Kremer (1996), Lakdawalla, Sood and Goldman (2006), Boucekkine, Diene and Azomahou (2008), Delavande and Kohler (2015), Gong (2015), Friedman (2018), and Greenwood et al. (2019). In the context of other recurrent diseases like the flu and malaria, some examples are Chakraborty, Papageorgiou and Pérez Sebastián (2010), Goenka and Liu (2012, 2019), and Goenka, Liu and Nguyen (2014). More recently, in the context of the COVID-19 pandemic, some examples are Acemoglu et al. (2020), Eichenbaum, Rebelo and Trabandt (2020*a*), Farboodi, Jarosch and Shimer (2020), Garibaldi, Moen and Pissarides (2020), Keppo et al. (2020), Krueger, Uhlig and Xie (2020), and Toxvaerd (2020). Similar to my paper, most of this literature build models in which agents can enjoy economic and social activity only at the expense of increased exposure to the virus. Thus, unsurprisingly, economic agents refrain themselves from many of the activities that they would pursue absent the epidemic, increasing the duration of the epidemic but significantly lowering life and welfare losses. My paper adds the importance of antibody testing in the context of the COVID-19 pandemic by showing the sizable gains from revealing to susceptible and asymptomatic agents that they are not immune.

## 2 Model

I build a simple economic model in discrete time with two blocks, an epidemiological block and a utility-maximization block.

### 2.1 Antibody Tests

Antibody tests are blood tests that aim to identify the presence of antibodies, which are disease-specific proteins that allow the body to fight infections and can provide immunity. As is common in the literature, in modeling antibody tests I assume that antibodies grant permanent immunity, implying that antibody tests identify recovered/immune agents.^5^

To model antibody tests, I divide agents in two groups that differ in health state uncertainty. A share *ω* of agents, whose variables are denoted by the superscript *τ*, continuously test their antibodies and, thus, *always* know whether they are immune; I think of these agents as having access to home antibody tests that can be used without medical supervision and with zero marginal cost;^6^ I refer to these agents as tested agents. The remaining 1 − *ω* share of the population, whose variables are denoted by the superscript *o*, never test their antibodies and, thus, *may* not know whether they are immune; I refer to these agents as untested agents.^7^ My assumptions imply that some untested agents are immune but, because they are unaware of it, they still behave as if they face exposure risk; furthermore, some untested susceptible agents may be excessively active because they think they may already be immune. My analysis will draw on how *ω* – governing the prevalence of antibody testing – affects the welfare loss and propagation of the epidemic.

In the model, I assume that antibody tests do not detect antibodies in infected agents. Even though antibody tests may theoretically identify a current infection (because of an early response of the immune system to the virus causing COVID-19), the usage of antibody tests to identify infected agents is not recommended by, e.g., the CDC (Centers for Disease Control and Prevention). Thus, for simplicity, I rule out this possibility; in the model, antibody tests only identify recovered agents. Furthermore, in this section, I abstract from imperfect testing and assume that antibody tests perfectly identify those that are immune and those that are not immune.^8^ Then, in Section 5, I adapt the model to assess the implications of imperfect antibody testing. Finally, I do not distinguish between different types of antibodies (namely IgA, IgM, and IgG). I simply treat antibody tests as a tool to identify immunity.

### 2.2 Epidemiological Block

The epidemiological block extends the canonical SIR (Susceptible-Infected-Recovered, Kermack and McKendrick, 1927) model. An agent can be in one of six health states: susceptible, *S*, asymptomatic, *E*, symptomatic, *I*, recovered undocumented, 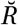, recovered documented, *R*, and dead, *D*. I denote the number of agents in group *y* = {*τ, o*} in state 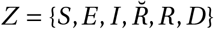 at time *t* by 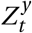. I also normalize the size of the population to unity, implying that 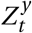 corresponds to both the number and the aggregate share of agents in each group and health state.

Infections in the model are governed by three factors: (i) the number of susceptible and infected (both asymptomatic and symptomatic) agents; (ii) *β* (a measure of the virus’ contagiousness); and (iii) agents’ social activity,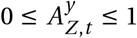 As a result, new infections of economic agents in group *y* at time *t* is 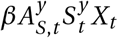 where *X*_t_ denotes all the social activity of infected agents (divided by the size of the population, *N*_*t*_):

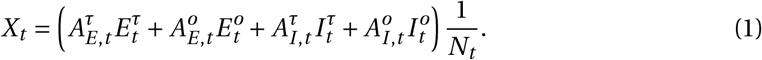

New infections start as asymptomatic, which is a state that lasts an average of 1/*γ*_*e*_ periods. Then, a proportion *σ* develop symptoms while the rest recover without developing symptoms and move to 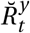 (recovered undocumented). Economic agents that develop symptoms remain infected for an additional 1/*γ*_*i*_ periods on average. And when they leave this state, a proportion *π* dies while the rest recover and move to 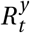 (recovered documented). As mentioned above, all recovered individuals develop antibodies and gain permanent immunity. The transitions between all states are summarized by:

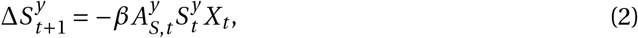

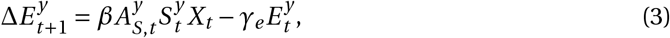

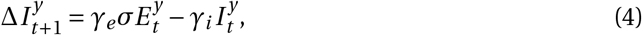

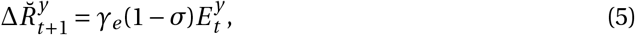

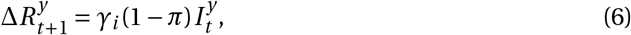

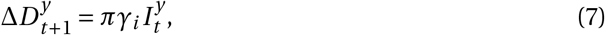

and population evolves according to: 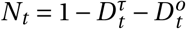.

I assume that the aggregate share of agents in each group and the degree of antibody testing is known.^9^ But agents do not perfectly know their own health state. In particular, I assume that symptomatic agents know that they are infected while asymptomatic agents do not know.^10^ When symptomatic agents recover, they are aware of the recovery but recovered undocumented agents are not as they were asymptomatic. Thus, I use the two recovered states to distinguish those who know that they are recovered (because they developed symptoms before) from those who do not know. Then, antibody tests create a wedge between the two groups of agents: all agents know whether they are recovered documented but only tested agents know whether they are recovered undocumented.

### 2.3 Agents’ Utility Maximization

The utility-maximization block of the model is close to that in Farboodi, Jarosch and Shimer (2020), Garibaldi, Moen and Pissarides (2020), Keppo et al. (2020), and Toxvaerd (2020) as agents directly choose social activity. There are, however, important distinctions. First, in Garibaldi, Moen and Pissarides, Keppo et al., and Toxvaerd, agents are always aware of their health state while in Farboodi, Jarosch and Shimer they are only aware of their health state when recovered. In my model, some agents know whether they are infected and not all agents know whether they are recovered. Second, their models assume homogeneous agents whereas I consider two groups of agents with access to different information. These nuances lead to different optimization problems, which allow me to study the implications of antibody testing.

In what follows, I use capital letters to denote aggregate variables and small letters to denote the variables of a specific agent. Thus, 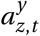 denotes the social activity of one agent in group *y* = {τ, *o*} in state 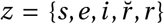 at time *t*. And 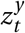 denotes the (subjective) probability of an agent in group *y* of being in state *z* at time *t*. Social activity *a* generates utility *u*(*a*); *u*(*a*) is strictly concave and continuously differentiable, has a maximum at *a*\* = 1 with *u*(*a*\*) = *u*′(*a*\*) = 0, and lim_*a*→0_+ *u*(*a*) = −∞. Moreover, each agent has rational beliefs about the probabilities of being in each health state and knows the transition probabilities between health states and how his or her social activity affects exposure to the virus. Each agent behaves atomistically, taking the evolution of *X*_*t*_ as given.

#### 2.3.1 Tested Economic Agents

Antibody tests partially reveal the health state: if the test is positive, agents learn that they are immune; if negative, agents learn that they are susceptible or asymptomatic (recall that symptomatic agents are aware of the infection). Thus, tested agents distinguish among all states except between susceptible and asymptomatic; when in these two states, they choose the same social activity: 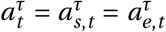. When tested agents are immune (i.e., recover from the infection), they optimally stop restraining their social activity; in this case,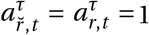. If (infected) symptomatic, I assume that the government imposes a quarantine: 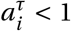, implying a per-period cost of being symptomatic.

The decision problem of tested economic agents is

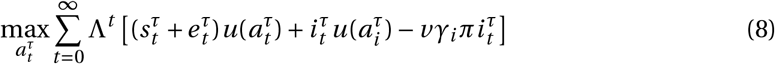

subject to

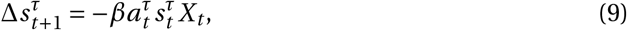

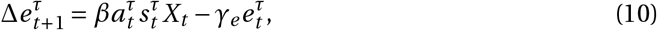

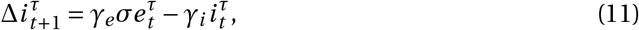

with 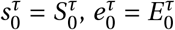, and 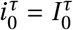 given. Using *v* to denote the value of life, the term 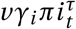 forces agents to account for the cost of dying (e.g., Farboodi, Jarosch and Shimer, 2020).^11^ 0 < Λ < 1 denotes the discount factor. The optimal behavior of agents with access to antibody tests is then determined by

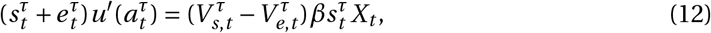

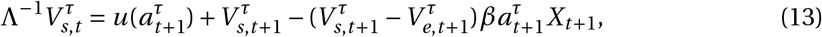

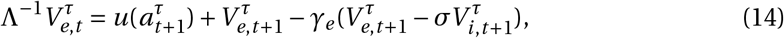

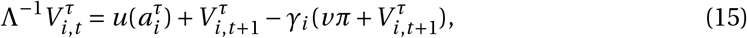

where 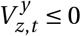 is the (shadow) value of agents in group *y* = {*τ, o*} of being in state 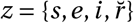 (see Appendix A.1 for more details). Because 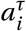 is fixed, the value of being symptomatic, 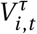 is constant and negative due to the imposed social-distancing and the risk of dying.

Eqs. (12-15) are all necessary conditions for optimality. To these equations, I add the three transversality conditions:^12^

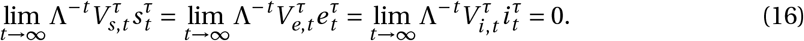

Eq. (12) summarizes the trade-off. Optimal behavior implies that the marginal gain from social activity (which is the marginal utility of social activity weighted by the probability of being susceptible and asymptomatic) equals the marginal cost (which depends on *β*, the shadow values, and the total social activity of infected agents, *X*_*t*_, and is weighted by the probability of being susceptible). Furthermore, because I restrict my analysis to the cases in which the value of being susceptible, 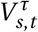, exceeds that of being asymptomatic,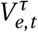, (as in Garibaldi, Moen and Pissarides, 2020) agents optimally reduce social activity to prevent exposure. As shown by Farboodi, Jarosch and Shimer (2020) this decentralized response of economic agents critically lessens the costs and the propagation of the pandemic. And it concurs with their evidence and the evidence reported in, e.g., Kaplan, Moll and Violante (2020) and Maloney and Taskin (2020) of behavioral changes in the US prior to lockdown policies.

#### 2.3.2 Untested Economic Agents

The maximization problem of tested and untested agents is similar. They only differ because untested agents do not distinguish the state of recovered undocumented from the states of susceptible and asymptomatic. Thus, social activity is the same when in these three states: 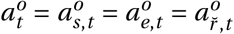. Untested agents also choose maximum social activity when recovered documented, 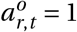, and their social activity is restricted when symptomatic 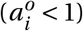. Thus, the decision problem of untested economic agents is

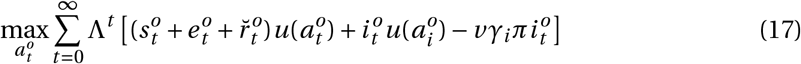

subject to

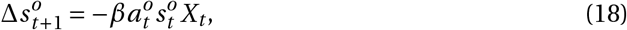

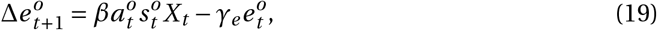

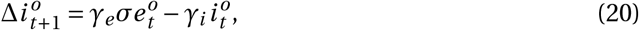

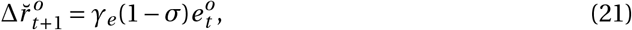

with 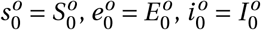, and 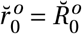 given. The optimal behavior of these agents is then determined by the following five necessary conditions (see Appendix A.2 for more details),

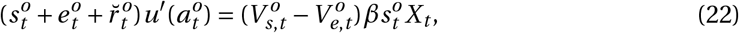

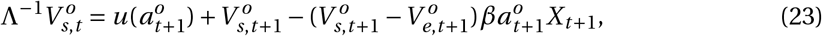

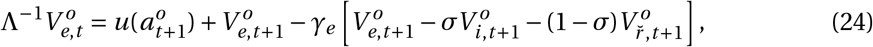

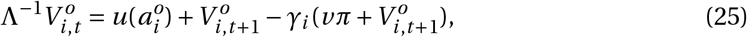

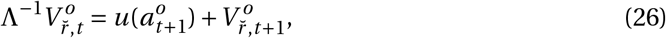

and the four transversality conditions,

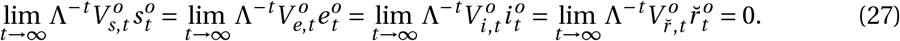

Information asymmetry generates three differences between the set of equations governing the optimal behavior of tested and untested agents. First, untested agents keep track of the value of being recovered undocumented, Eq. (26), whereas tested agents know that their value in this state is always the maximum possible (normalized to zero). Second, comparing Eqs. (12) and (22) shows that untested agents attach, *ceteris paribus*, a higher weight to the marginal utility of social activity than tested agents. This motivates untested agents to raise social activity (above that of tested agents), especially when 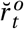 is large. Intuitively, when 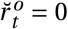, agents are more cautious as they are likely susceptible and, thus, may be exposed to the virus. But when 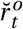 is large, untested agents *think* they might be immune; as constraining social activity is unwarranted when immune and there is health state uncertainty, untested susceptible and asymptomatic agents prefer to enjoy more social activity and take more risks. Third, comparing Eqs. (14) and (24) shows that the (negative) value of being untested recovered undocumented reduces, *ceteris paribus*, the value of being untested asymptomatic. This, in turn, raises the right-hand side of Eq. (22), which tends to reduce the social activity of untested agents. Intuitively, as untested agents anticipate the possibility and the costs of being recovered undocumented, they prefer to restrict their social activity (relative to tested agents) to reduce the probability of being in that state. Yet, in my simulations, this last force plays a minor role and untested agents end up choosing higher social activity than tested agents when 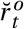 is slightly positive.

### 2.4 Equilibrium

All agents within each group have the same preferences and access to the same information. Thus, at all *t*, their social activity,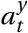 and 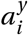, equals aggregate social activity, 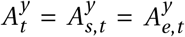 and 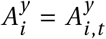. This implies that the probability of being in each state must equal the population share in that state: 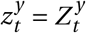 at all *t*.

Given initial values for the state variables, a decentralized equilibrium satisfies the transversality conditions (Eqs. 16 and 27) and corresponds to a path of social activities, 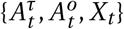, state variables, 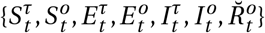, and shadow values, 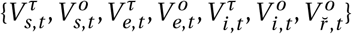, that satisfy Eqs. (1-5), (12-15), and (22-26).

## 3 Calibration

I calibrate the model to daily data and summarize my parameter choices in Table 1. I set 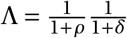, where *ρ* = 0.05/365 is the time discount rate and *δ* = 0.67/365 is the probability of finding a cure/vaccine (e.g., Alvarez, Argente and Lippi, 2020; Farboodi, Jarosch and Shimer,2020).

**Table 1:**
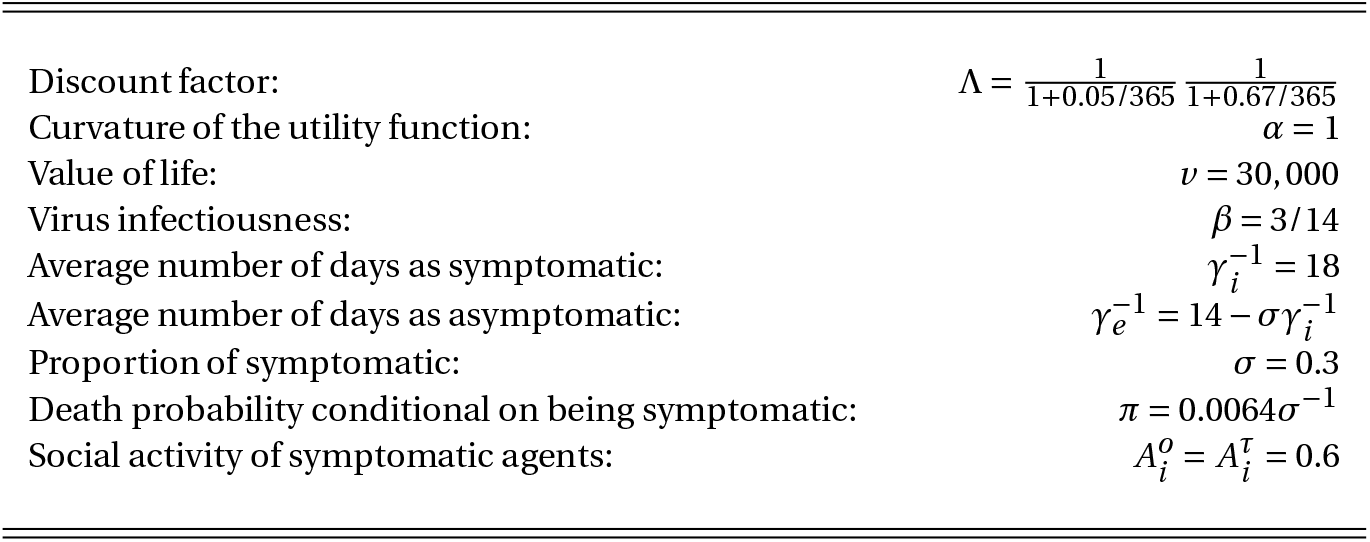
Benchmark Calibration.

I assume the following functional form for the utility of social activity:

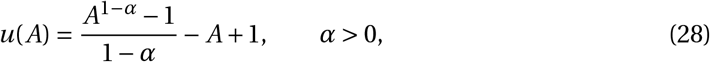

which guarantees that *u*(*a*) is single-peaked with maximum at *A* = 1 and *u*(1) = *u*′(1) = 0. As my benchmark, I set *α* = 1, which delivers the same utility function as in Farboodi, Jarosch and Shimer. I also closely follow Farboodi, Jarosch and Shimer to find the value of life, *v*, in model units. I start by using a value of a prevented fatality in the UK of £1.8 Million (Thomas and Waddington, 2017) and by interpreting this number as agents being willing to pay £1.8 thousand to avoid a death probability of 0.1% or, equivalently, £0.247 per day (using the yearly discount rate of 5%). Given UK consumption per capita of £22.2*k*, this implies that agents are willing to permanently trade 0.41% of their consumption for a permanent reduction in their death probability of 0.1%. Assuming that the utility of consumption refers to the first term on the right-hand side of Eq. (28) (and using *α* = 1) implies that the value of life must satisfy the following indifference condition:

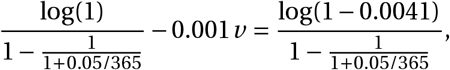

and, thus, *v* = 30, 000. This is lower than the value found by Farboodi, Jarosch and Shimer because I assume a relatively low value of prevented fatality.

I follow Atkeson (2020) and Wang et al. (2020) and assume that agents remain infected for 18 days after developing symptoms: 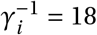. To find *γ*_*e*_, I target an average infection time of 14 days, which is the WHO guideline for the duration of COVID-19-related quarantines. Given that individuals stay asymptomatic for 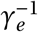 periods and a proportion *σ* of them remain infected (but symptomatic) for an additional 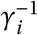 days, I obtain *γ*_*e*_ using 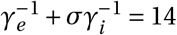.

Most of the infected agents recover after infection but some die. Meyerowitz-Katz and Merone (2020) conduct a meta-analysis of published estimates of COVID-19 infection-fatality rates and conclude that about 0.64% of those infected with the virus die. Thus, I fix *π* = 0.0064/*σ*, implying that slightly more than 2% of symptomatic agents die due to the infection.

In epidemiological models, the *basic* reproduction number, *R*0, is key to determine the number of infections and herd immunity to a virus. In the model, given that agents remain infected for an average of 14 days, I set *β* = 3/14. This implies that *R*0 = 3 absent quarantining of symptomatic agents and behavioral responses to exposure risk, which is close to the numbers typically used in the economics and epidemiological literatures.

I fix *σ* = 0.3, implying that 70% of all infected agents do not develop symptoms or, at least, do not have access to a viral test confirming the infection. *σ* = 0.3 is close to the lower bound (0.15) of the early estimates reviewed in Eikenberry et al. (2020) and Stock (2020). *σ* = 0.3 also agrees with the proportion of estimated asymptomatic individuals in the UK based on the ONS (Office for National Statistics) survey.^13^ The estimated share of asymptomatic in Spain is lower (around 33%; see Pollán et al., 2020). Yet, given that I assume that symptomatic agents are sure of being infected, I also look at the proportion of those with antibodies that were diagnosed with the infection. The ONS survey suggests that, in June 2020, about 5.4% of the UK population had developed antibodies against COVID-19 but less than 0.5% of the population was diagnosed with an infection. In Spain, comparable numbers are 5% and 0.6%, respectively. These numbers suggest that *σ* = 0.3 is reasonable.

Finally, I follow the calibrations of lockdown measures and lockdown effectiveness in Acemoglu et al. (2020) and Alvarez, Argente and Lippi (2020) to set 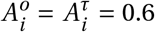. To solve the model, I use a shooting algorithm as detailed in Garibaldi, Moen and Pissarides (2020) and assume that initially one in a million individuals are asymptomatic while the rest is susceptible.

I do not impose a value for the proportion of untested agents, *ω*. My objective is to test for the interval of *ω* ∈ [0, 1] to see how different values affect the results. I also conduct several robustness checks to changes in the parameters of the model.

## 4 Results

### 4.1 Main Results

Figure 1 summarizes how the the share of tested agents, *ω*, affects three indicators of the effects of an epidemic under the benchmark calibration. The model suggests that, if all agents are continuously tested (*ω* = 1), 50% of the population is exposed to the virus within one year, which contrasts with 56.7% if no agent is ever tested (*ω* = 0). In other words, widespread and continuous antibody testing reduces the number of exposed individuals and, thus, deaths by 11.8%. Antibody testing also rises welfare substantially for both tested and untested agents because it reduces exposure to the virus, life losses, and increases social activity of immune agents.

**Figure 1:**
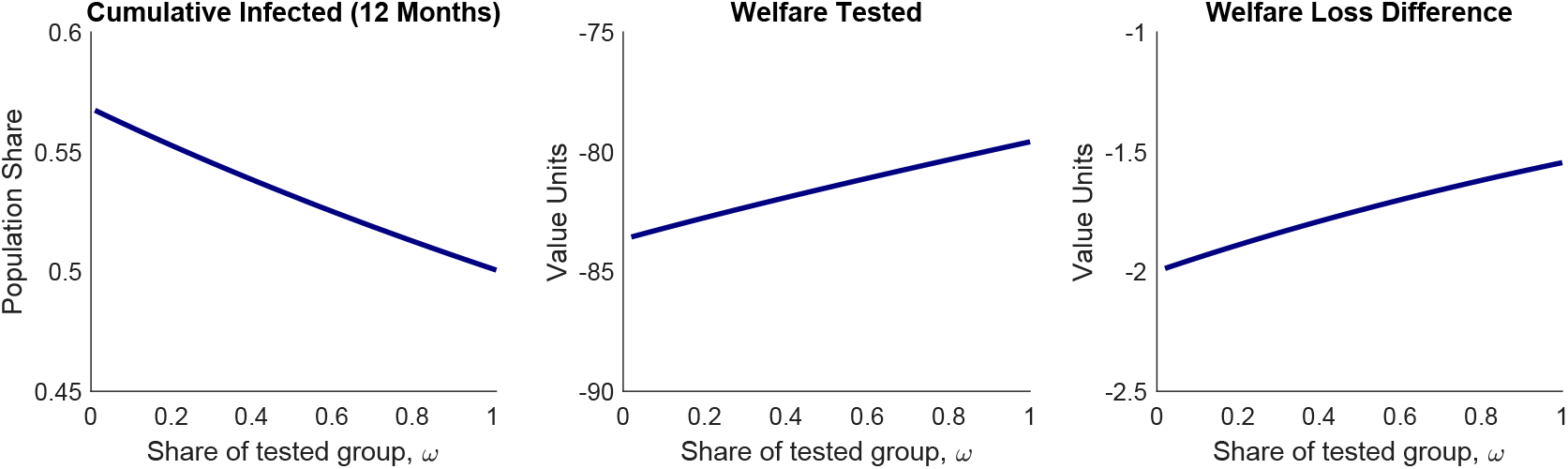
Main Results. *Note:* This figure shows cumulative infections after one year, 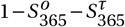, welfare of tested susceptible agents in period 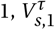, and difference in welfare loss between tested and untested susceptible agents in period 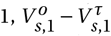, as a function of the share of tested agents, *ω*. The model is calibrated using the benchmark calibration.

To understand why antibody testing reduces total exposure and welfare losses, Figure 2 contrasts the social activity of tested agents with that of untested agents when half of the population is tested (*ω* = 0.5) and under multiple calibrations of the model. The key takeaway from this figure is that tested agents restrain their social activity by more than untested agents, *A*^*τ*^ < *A*^*o*^, especially about 100 days after the beginning of the epidemic (in the benchmark).

**Figure 2:**
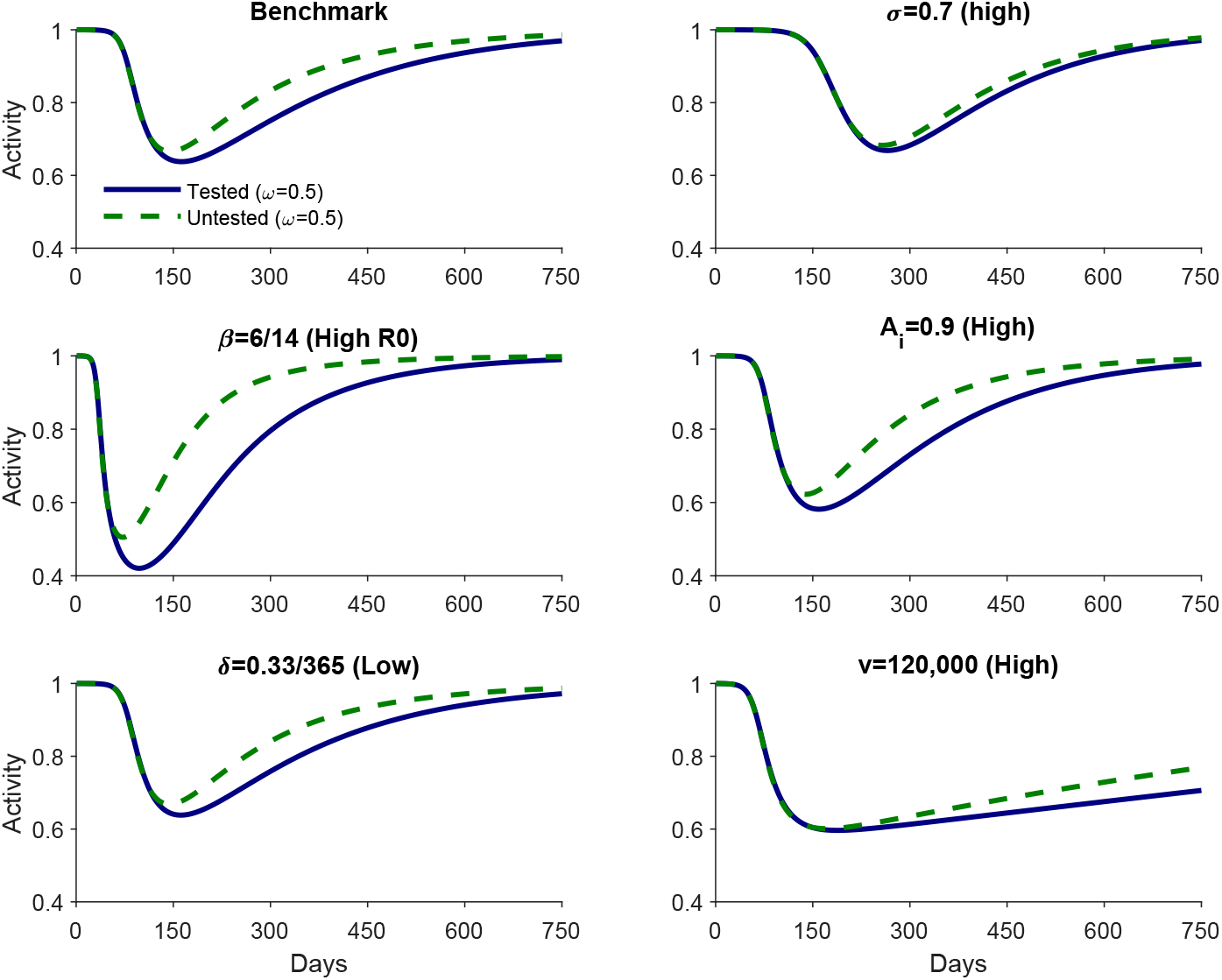
Social Activity Under Different Calibrations. *Note:* This figure shows social activity of tested and untested susceptible agents under six calibrations of the model and assuming that half the population is tested, *ω* = 0.5.

The reason was already hinted in Section 2.3: as the probability of being recovered undocumented enlarges, untested susceptible and asymptomatic agents start increasing social activity with the hope that they are immune but are unaware of it. Therefore, testing agents lowers the average social activity of susceptible agents and, very importantly, the total social activity of infected agents, *X*_*t*_; this implies, respectively, a direct and an *externality* effect reducing total exposure to the virus and the welfare losses of all agents.

Figure 1 also shows that untested agents suffer more from the epidemic than tested agents (Welfare Loss Difference is negative). Even though untested agents enjoy more social activity, the losses from the higher exposure to the virus escalate their losses. Yet, somewhat surprisingly, antibody testing increases the welfare of untested agents by more than that of tested agents (the slope of the Welfare Loss Difference curves is positive). In other words, untested agents benefit marginally more than tested agents if there is an increase in the share of tested agents, *ω*. The cause is, once more, the *externality* effect of antibody testing on *X*_*t*_, which reduces exposure of all susceptible agents to the virus and allows them to enjoy more social activity.^14^ Because 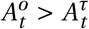, the fall in *X*_*t*_ is especially important to reduce exposure of untested agents.

To sum up, by increasing the information available to economic agents, antibody testing lowers contagion, increases welfare, and reduces inequality. The reason is simple: susceptible and asymptomatic agents who know that they are not immune are less socially active.

### 4.2 Robustness Checks

In this section, I show that my qualitative results are robust to different calibrations but also that some parameters substantially change the quantitative results as indicated by the slopes of the lines.

The green dashed lines in Panel A of Figure 3 present the case of *σ* = 0.7, i.e., 70% of asymptomatic individuals eventually become symptomatic. This experiment suggests that the gains from antibody testing depend negatively on *σ* because a higher share of recovered individuals know that they are immune even without antibody tests. It also suggests that large-scale viral testing and efficient contact-tracing (to detect asymptomatic agents early) narrow the role of antibody testing in revealing information.

**Figure 3:**
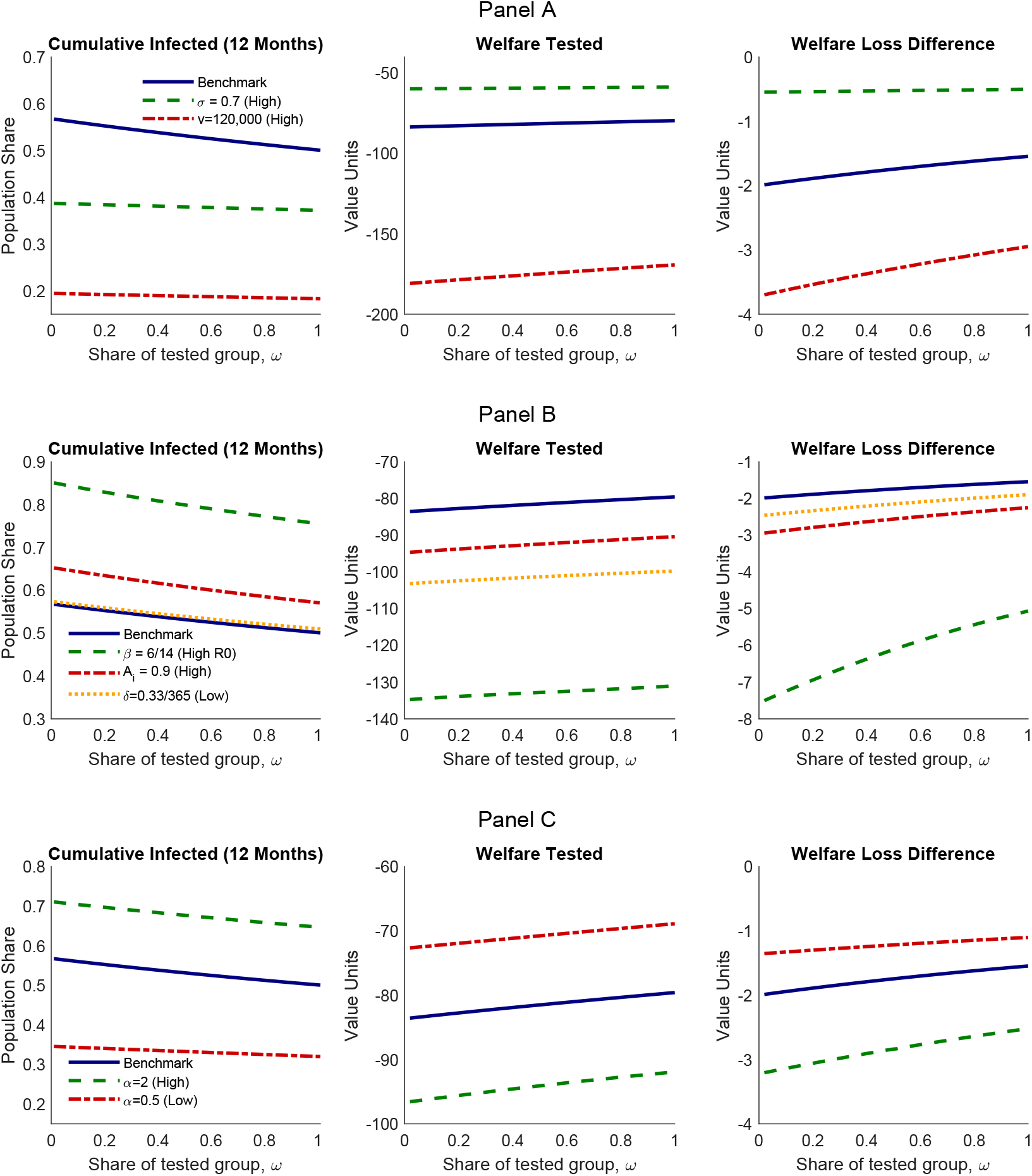
Robustness Checks. *Note:* See the note to Figure 1 for details about the variables.

The value of a prevented fatality that I use to calibrate *v* is relatively low. It is slightly higher than in Alvarez, Argente and Lippi (2020) and slightly lower than in Hall, Jones and Klenow (2020) but much lower than in, e.g., Eichenbaum, Rebelo and Trabandt (2020*a,b*) and Farboodi, Jarosch and Shimer (2020). The red dot-dashed lines in Panel A of Figure 3 show the implications of *v* = 120, 000 (four times larger than my benchmark). If *v* = 120, 000, the number of exposed agents fall approximately 64% relative to benchmark because all economic agents persistently constrain their social activity (see Figure 2). But, importantly, the welfare gains of antibody testing are still substantial: even though antibody testing prevents less deaths, the value attached to each life is four times higher.

I also experimented with an extreme basic reproduction number of *R*0 = 6 and report the results using the green dashed lines in Panel B of Figure 3. In this case, the (absolute) slope of all lines increases, suggesting that the more contagious is the virus, the more important are antibody tests. By the same token, if NPIs like mandatory mask use reduce contagiousness, they also reduce the effectiveness of antibody testing.

Figure 3 also reports the consequences of lenient quarantines of symptomatic agents, 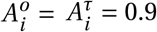, and a low probability of finding a cure/vaccine, *δ* = 0.33/365, in Panel B and a different curvature of the utility function, *α*, in Panel C. Most of these experiments show major effects on the propagation of the epidemic. But there are not significant changes in the slopes of the lines.

### 4.3 Long-Run Effects of Antibody Testing

My analysis so far has focused on the short-run (within the first year) gains from antibody testing. But, as a vaccine or cure may not arrive, there are natural concerns about the longrun effects of antibody testing. In particular, does it reduce long-run cumulative infections or merely postpones exposure to the virus? To answer this question, Figure 4 reports a phase diagram for the evolution of the pandemic from its starting point until it disappears in the long run.^15^ The phase diagram shows the combinations of susceptible agents 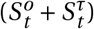 and infected agents (both asymptomatic and symptomatic, 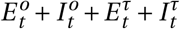 over time. The pandemic starts on the bottom-right of Figure 4 where there are very few infected agents and almost all agents are susceptible; it continues as contagion leads to further infections and agents lose their susceptibility to the virus; asymptotically, it ends on the bottom left as the number of new infections continuously drop because of the fall in susceptible agents.

**Figure 4:**
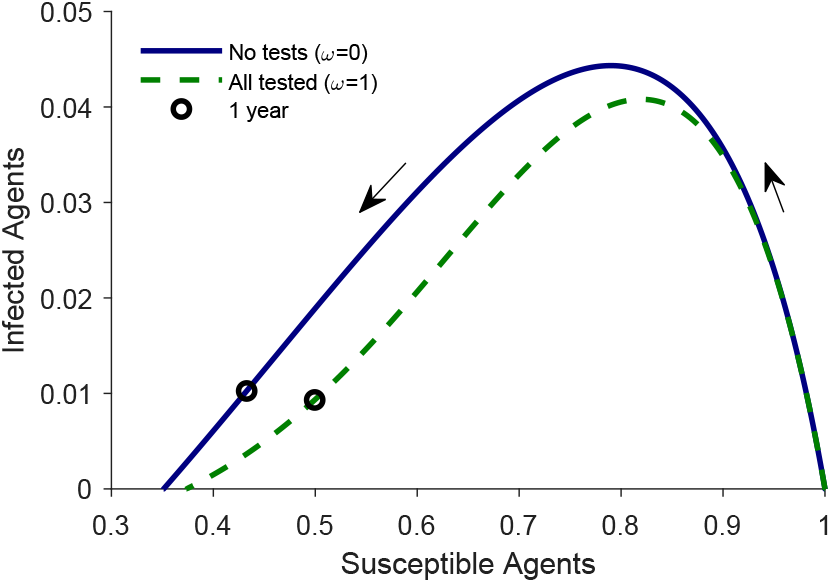
Disease Dynamics. *Note:* This figure shows the combinations of susceptible agents 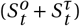 and infected agents (both asymptomatic and symptomatic, 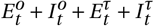 over time. The black circles identify the circumstances after one year.

To show the gains from antibody testing, Figure 4 plots the two extremes: *ω* = 0 (no agent is ever tested) in the blue solid line and *ω* = 1 (all agents are continuously tested) in the green dashed line. The solid line is repeatedly above the dashed line, implying that there are more infections for each level of susceptible agents in the scenario without antibody tests. This confirms that antibody testing persistently reduces infections and, thus, COVID-19 related deaths. Figure 4 also shows that the distance between the two lines narrows asymptotically, suggesting that one of the main benefits of antibody testing is to postpone infections and gain time for a vaccine or cure to arrive. But, even in the long run, thanks to antibody testing, cumulative infections (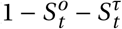 as *t* → ∞) shifts from 64.9% when *ω* = 0 to 62.6% when *ω* = 1.^16^ Put differently, in the long run, cumulative infections and COVID-19 related deaths are 3.5% lower due to antibody testing.

## 5 Single & Imperfect Testing

In this section, I study the scenario in which home antibody tests that can be used with zero marginal cost are not available and tested agents are tested only once at an unannounced date.^17^ This section complements the previous ones in two ways. First, as there is the risk that home antibody tests are not widely available in a timely manner, this section complements the previous ones by grasping the importance of large-scale antibody testing using tests already widely available. Second, as agents are tested only once, the problems caused by imperfect testing can be highly relevant. Current antibody tests are imperfect as some immune agents test negative for antibodies against the virus causing COVID-19 while some susceptible agents test positive. This imperfection naturally reduces the ability of antibody tests to inform economic agents, which lowers their effect on contagion and lives saved. Therefore, this section also complements the previous ones by assessing the consequences of imperfect testing.

### 5.1 Model

Tested and untested agents hold the same information and behave alike until the day of the test. But after the test, tested agents update their subjective probabilities depending on the test result and start behaving differently. This implies a split between tested and untested agents but also among tested agents as the test conveys different information depending on the test result. Therefore, I split tested agents in two groups, those that test positive and those that test negative. Using the superscripts *pτ* and *nτ* to denote agents that test (respectively) positive and negative and using *p* and *q* to denote false positive and false negative rates, tested agents are distributed according to:

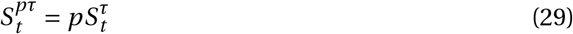

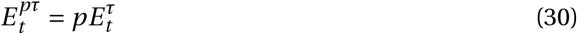

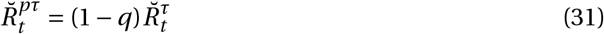

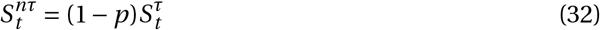

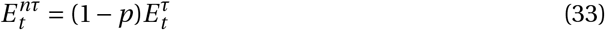

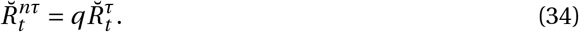

Infected and recovered documented agents are unaffected by the test as they know for sure that they are infected and recovered, respectively. If *p* = *q* = 0, tests are perfect and agents know with certainty whether they are immune. If *p* = 0 and *q* > 0, then those that test positive are sure to be immune, while if *p* > 0 and *q* = 0, those that test negative are sure to not be immune.

After the test, tested agents have rational beliefs about the probability of being in each health state based on their test results. Furthermore, tested agents know that they will never be tested again. Thus, after the test, tested agents face the same problem as untested agents, which is detailed in Section 2.3.2. The only difference between untested agents and those that test positive and negative regards their subjective probability of being in each health state, which I assume to initially equal the respective aggregate share of the population. As those that test positive have a higher probability of being recovered undocumented, they tend to be more active than other agents.

### 5.2 Results

In producing the results in this section, I assume that all agents are tested, i.e., *ω* = 1. Thus, my experiments in this section lie between the extremes of no testing and continuous and widespread testing in the previous section.

Table 2 shows how the test day and the degree of imperfect testing change the cumulative exposure to the virus in the first 12 months and in the long run. As a benchmark, I consider the case of no testing (*ω* = 0); thus, Table 2 reports the percentage reduction in cumulative infections caused by antibody testing relative to the scenario without tests.

**Table 2:**
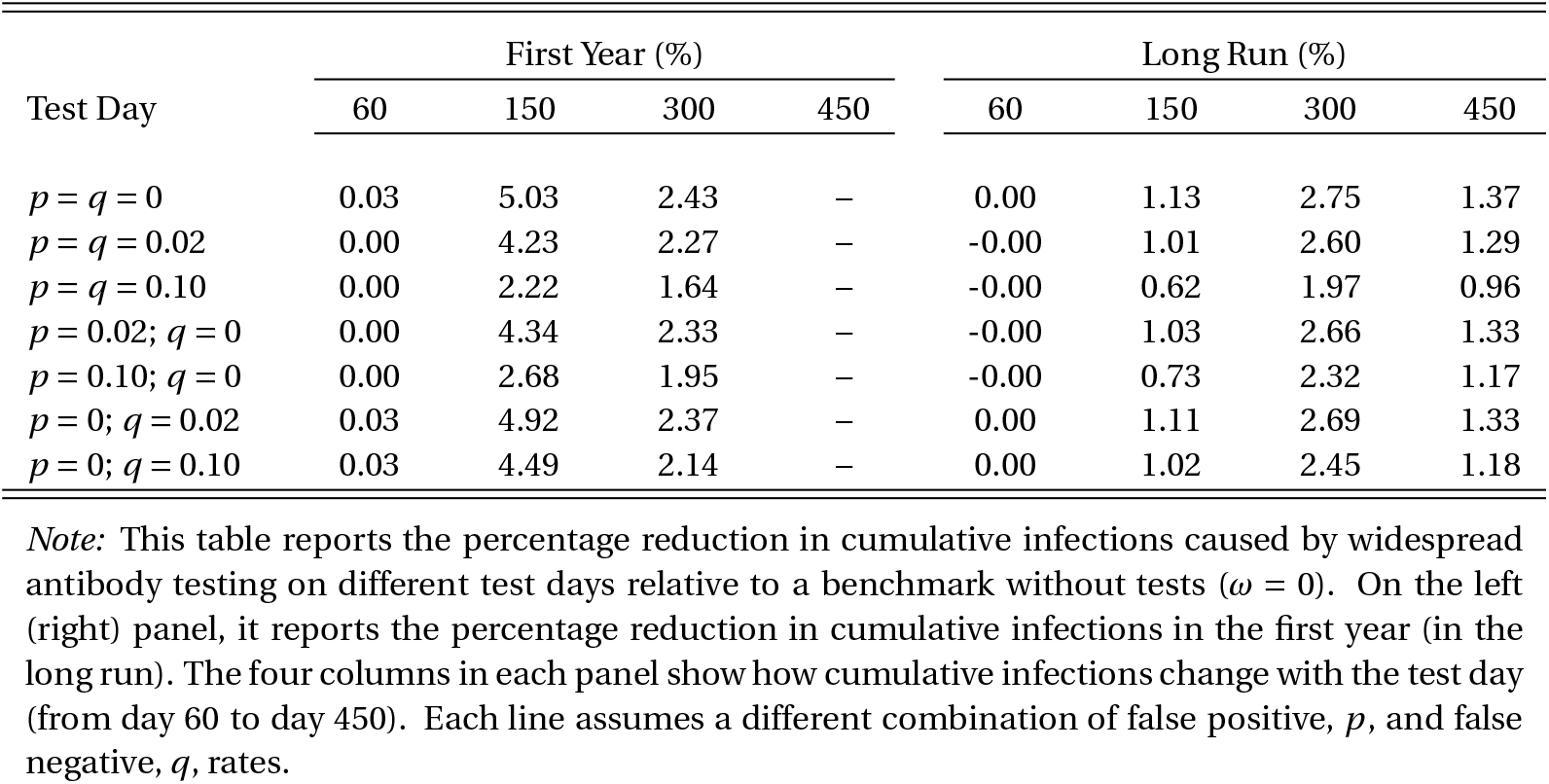
Benefits of Single & Imperfect Testing - Reduction in Cumulative Infected.

Table 2 shows that widespread antibody testing, even if not continuous, significantly constrains contagion and saves lives. For example, compared with no testing, testing all agents five months past the beginning of the pandemic reduces cumulative infections in the first 12 months by 5%, which is is almost half of that obtained if agents are continuously tested (11.8%). Panel B in Figure 5 helps to explain this result: under perfect testing, agents with a negative test learn that they are not immune and permanently lower their subjective probability of being recovered undocumented; therefore, these agents significantly and persistently reduce their activity, lessening the extent of the pandemic.

**Figure 5:**
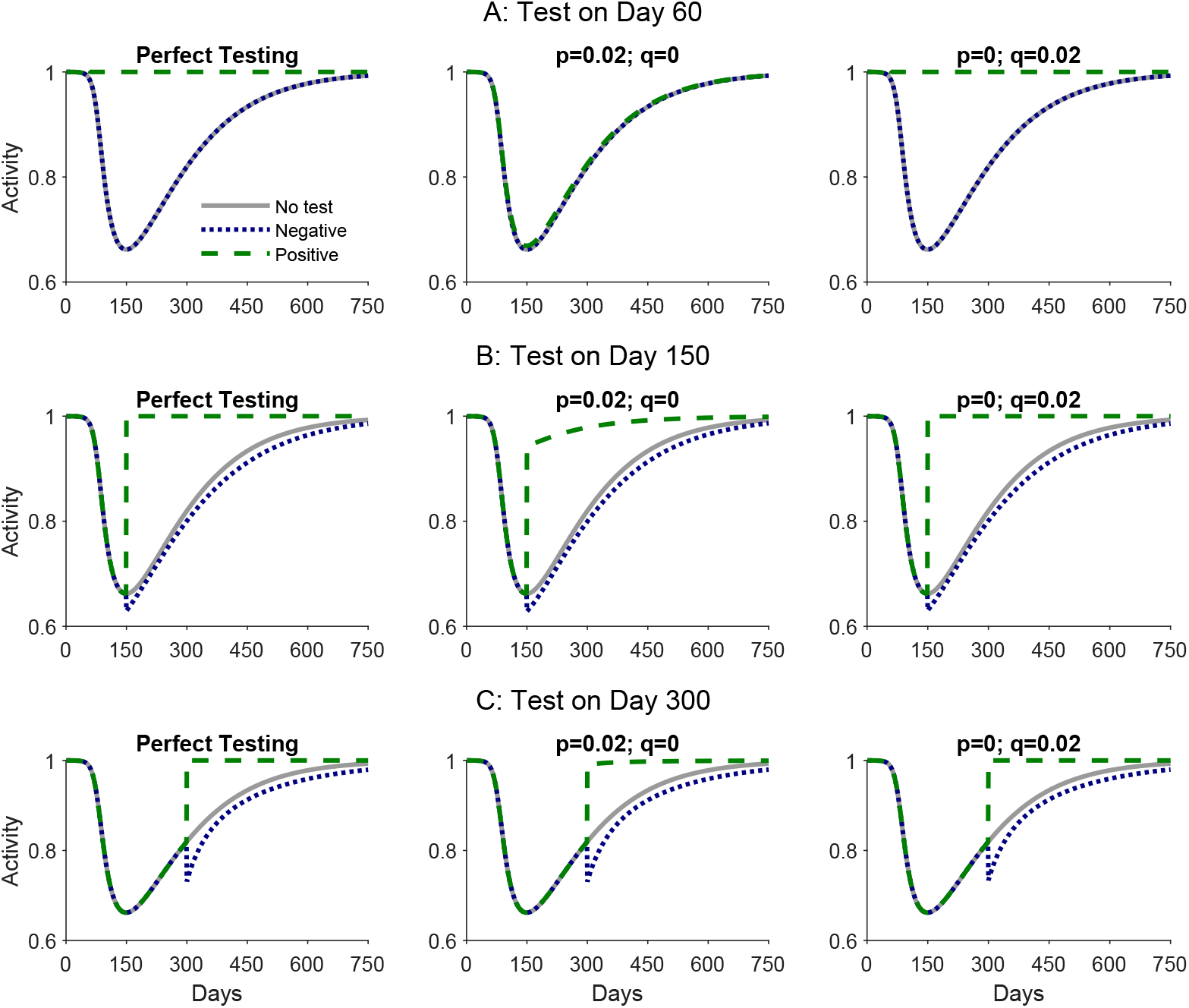
Single & Imperfect Testing - Social Activity. *Note:* This figure contrasts the social activity in the scenario without tests (*ω* = 0) with the social activity of those that test positive and test negative on a particular test day and under different false positive, *p*, and false negative, *q*, rates.

Table 2 also highlights the importance of the test day.^18^ As hinted by the results in the previous section, antibody testing occurring early in the pandemic (e.g., on day 60) barely affects the disease dynamics because very few agents are recovered undocumented. Panel A in Figure 5 confirms this logic because, even under perfect testing, there are no visible differences in the optimal social activity of agents that obtain a negative result relative to the benchmark without tests. Furthermore, Table 2 shows that it is less fruitful to test at a late stage (e.g., on day 450) because much of the disease dynamics have already taken place. Yet, it also shows that testing later in the pandemic can have more long-lasting effects on cumulative infections. Contrasting Panels B and C in Figure 5 shows that tests on day 150 allow for an earlier reduction in activity, which result in an earlier reduction in infections. But tests on day 300 lead to a larger hiatus in activity as they resolve more health state uncertainty.^19^ This, in turn, leads to a substantial reduction in exposure on the final stages of the pandemic.

To assess the implications of imperfect testing, I report six further combinations of false positive, *p*, and false negative, *q*, rates in Table 2. These combinations are based on the performance of various antibody (serology) tests reported by the US Food and Drug Administration.^20^ In most cases, the estimated sensitivity and specificity are very large and close to 100%. Yet, as the number of samples is small, the confidence interval of those estimates is wide implying false positive and false negative rates ranging from 0% to, in some cases, more than 10%. Hence, in my experiments I consider 0 ≤ *p* ≤ 0.1 and 0 ≤ *q* ≤ 0.1.

I find that imperfect testing significantly diminishes the gains from antibody testing. For example, if *p* = *q* = 0.1 and the test occurs on day 150, then antibody testing reduces cumulative infections in the first 12 months by 2.2% instead of 5% with perfect testing. Most of this reduction in the benefits of antibody testing is driven by the false positive rate. And the panels in the middle column in Figure 5 help to shed light on the reason. If *p* > 0, some susceptible and asymptomatic agents obtain a false positive result. Accordingly, as there is the risk of being susceptible even with a positive test, all agents with a positive test are forced to still constrain their activity. Yet, on day 150 or later, an agent with a positive test is most likely recovered undocumented; thus, susceptible and asymptomatic agents that obtain a positive test become more active, leading to a lower reduction in infections than otherwise.

False negative tests also curb down the benefits of antibody testing in terms of reducing the social activity of susceptible and asymptomatic agents. If there are multiple false negatives, then the test is less informative and susceptible and asymptomatic agents that obtain a negative result barely react to it. I find, however, that this effect, though relevant, is less quantitatively important even if *q* = 0.1.

I also find that the effects of antibody testing on welfare depend on the health state of the agent and on the test result (see Tables B1-B3 in the Appendix). Unequivocally, asymptomatic agents benefit from a positive antibody test and lose from a negative antibody test as they are not altruistic and, thus, prefer unconstrained activity. A similar result applies to recovered agents, who would also prefer to not constrain their activity as they cannot be reinfected. Interestingly, the change in the welfare of recovered agents depends deeply on the degree of false positives. For example, an early and imperfect antibody test (e.g., on day 60) barely offers any information and, thus, barely increases welfare for these agents if positive.^21^ But antibody testing at a later stage significantly increases (lowers) their welfare if positive (negative). Finally, susceptible agents always benefit from a negative antibody test because, if there was perfect information, they would choose lower social activity to reduce exposure to the virus. Yet, the effect of a positive test on the welfare of susceptible agents depends on the test day. There are two effects. First, as susceptible agents become even more active after a positive test, their well-being worsens. Second, as many susceptible and asymptomatic agents restrict their activity after negative tests, the total social activity of infected agents, *X*_*t*_, tends to fall, leading to a general-equilibrium effect that reduces exposure of all susceptible agents. Surprisingly, this general-equilibrium effect is stronger if the test occurs in the later stages of the pandemic (e.g., on day 300 or later).

## 6 Concluding Remarks

In the context of the COVID-19 pandemic, the literature advocates many NPIs to reduce contagion and economic costs. Alvarez, Argente and Lippi (2020) and Glover et al. (2020) argue that lockdowns and quarantines can increase welfare; Acemoglu et al. (2020) and Gollier (2020) argue that age-specific lockdown and quarantine policies are far better than age-indifferent ones; Mitze et al. (2020) document that mandatory mask use reduces deaths; Berger, Herkenhoff and Mongey (2020), Brotherhood et al. (2020), Eichenbaum, Rebelo and Trabandt (2020*b*), and Piguillem and Shi (2020) argue that combining viral testing and quarantining increases welfare. In this paper, I argue that antibody testing is another NPI – not recognized so far in the literature – that reduces contagion. The point is that susceptible and asymptomatic agents that are unaware whether they are immune are more socially active than those who are sure of not being immune. Therefore, by increasing the number of agents who are sure of not being immune, antibody testing saves lives and increases welfare.

My model also hints that the welfare gains of antibody testing and its efficacy in preventing COVID-19 contagion depends:

- Positively on the contagiousness of the virus because the effects of increased activity in the absence of antibody tests compound quickly;
- Positively on the probability of being asymptomatic because it increases the probability that agents are unaware that they are immune;
- Negatively on viral testing and contact tracing because (i) these policies increase the number of individuals aware of recovery absent antibody tests and (ii) also reduce contagiousness;
- Negatively on other interventions that likely lower contagiousness like mandatory mask use.
- Negatively on the degree of imperfection in the antibody tests, especially if sensitivity falls short of 100%.

My simulations suggest that antibody testing becomes particularly important when a sufficiently large proportion of the population is unaware of its immunity. This is the case in countries like the US, UK, Spain, France, and other European countries but, even more clearly, in places like Delhi, London, Madrid, and New York where more than 15% of the population is estimated to be immune. Those numbers, although staggering, fall significantly short of herd immunity. Thus, my model flags the importance of identifying those who are not immune to lower the future propagation of COVID-19.

## Data Availability

All data is freely available.

## A Derivation of the Decision Problems

### A.1 Tested Agents

To solve the optimal control problem of tested agents set in Section 2.3.1, I start by writing the discounted Hamiltonian:

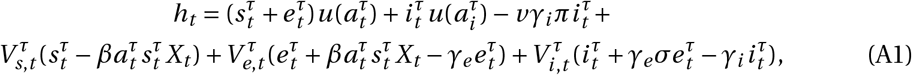

where 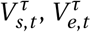, and 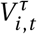 are the co-states. Using 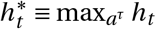, the optimal path must satisfy the transversality conditions and

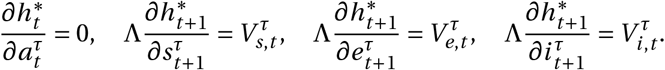

Solving yields Eqs. (12-15).

### A.2 Untested Agents

To solve the optimal control problem of untested agents set in Section 2.3.2, I again start by writing the discounted Hamiltonian:

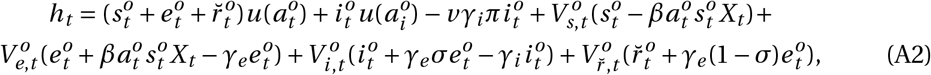

where 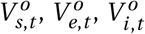, and 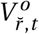 are the co-states. Using 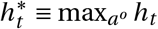, the optimal path must satisfy the transversality conditions and

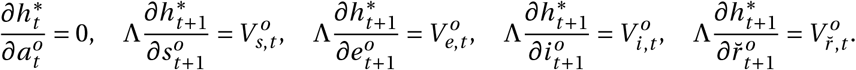

Solving yields Eqs. (22-26).

## B Sufficiency Conditions for Optimality

To determine whether the path implied by Eqs. (12-16) is optimal, I contrast the value of the Hamiltonian in Eq. (A1) under different paths of activity, 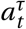. The benchmark is the path implied by Eqs. (12-16), which henceforth I refer to as optimal.

My approach in this appendix is numerical, which does not allow me to prove that the optimal path is indeed optimal but does suggest that it is. Importantly, as I only study decentralized equilibria, I look at the behavior of a marginal tested agent that is unable to affect aggregate variables (namely *X*_*t*_) and, thus, takes them as given.

### B.1 Steps

All my experiments follow these steps:

1. Calibrate the model according to Table 1. In producing the results in this appendix, I further assume that *ω* = 0.5.
2. Assume a given path for *X*_*t*_. In this appendix, given the initial conditions specified in Section 3, I consider two feasible paths of *X*_*t*_ in separate scenarios:
  a. In one scenario, I assume the (aggregate) equilibrium *X*_*t*_. In this scenario, the optimal path of the marginal agent equals that of the other agents with the same characteristics.
  b. In another scenario, I assume that all other agents are fully active in all periods, which is not the equilibrium. This scenario allows me to present results suggesting that the optimal path (for given *X*_*t*_) of a marginal agent is optimal even if the economy is not in the (aggregate) equilibrium.
3. Obtain the optimal path of 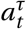 as implied by Eqs. (12-16) given *X*_*t*_ and the initial conditions specified in Section 3. Henceforth, I denote the optimal path of 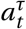 as 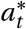. Based on 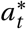, calculate the value of the state variables over time and of the shadow values of the marginal agent. Then, calculate the value of the Hamiltonian at *t* = 1, using Eq.(A1).
4. Consider alternative feasible paths of 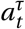. Given the initial conditions specified in Section 3, for each path, calculate the associated value of the state variables over time and of the shadow values of the marginal agent. Then, calculate the value of the Hamiltonian at *t* = 1, using Eq. (A1).
5. Finally, contrast the value of the Hamiltonian when the optimal path is followed and when another path is followed. This step offers a measure of loss caused by shifting from the optimal path to an alternative path; in particular, using 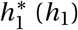 to denote the value of the Hamiltonian when the optimal (alternative) path is followed, I report 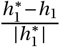.

### B.2 Experiments

I consider three experiments, one suggesting that the optimal path is locally optimal and the other two experiments suggesting that the optimal path is globally optimal.

To produce Figure B1, I add (left panel) or subtract (right panel) ϵ= 0.0001 to *a*\* for each *t* ∈ [1, 2000]. In all 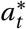, the loss is at least zero, suggesting that the optimal path is indeed locally optimal, irrespective of *X*_*t*_.

**Figure B1:**
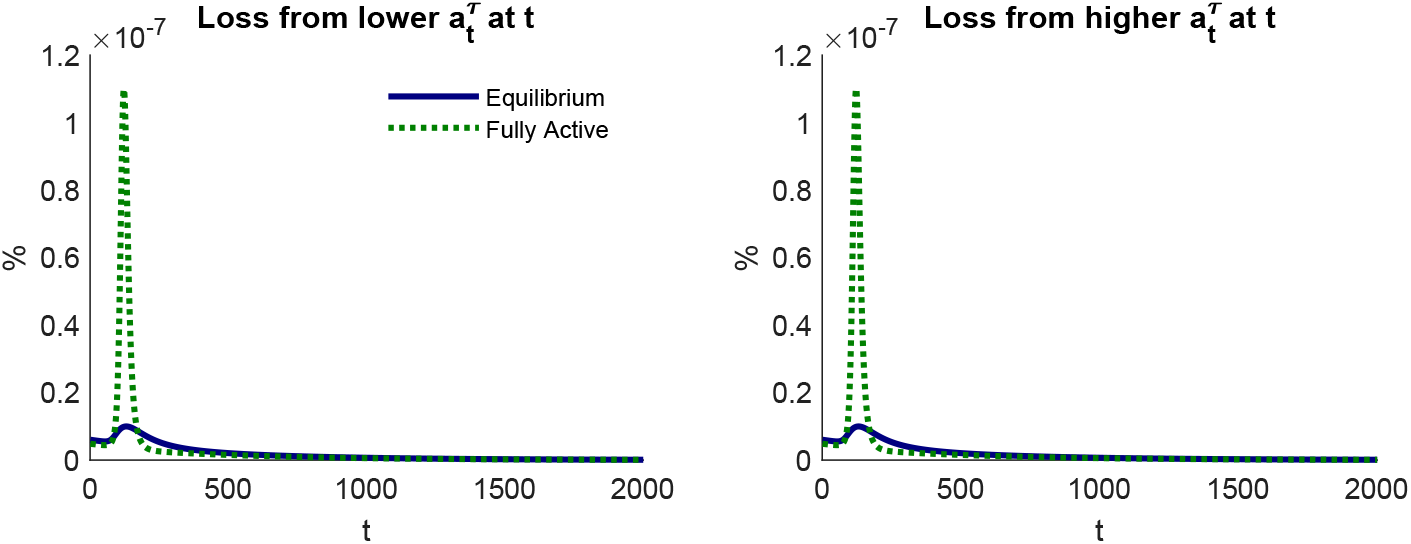
Local Optimality.

Figure B2 suggests that the optimal path is globally optimal. On the left panel, this figure shows the loss caused by deviating from 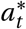 to 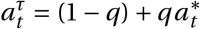, i.e., a weighted average of maximum activity and the optimal path. On the right panel, the alternative path is given by 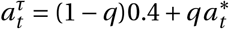, i.e., a weighted average of 0.4 (set arbitrarily low) and the optimal path. In all cases, the lower is *q* (the larger is the difference between the alternative path and the optimal path), the lower is the value of the Hamiltonian, irrespective of the path of *X*_*t*_.

**Figure B2:**
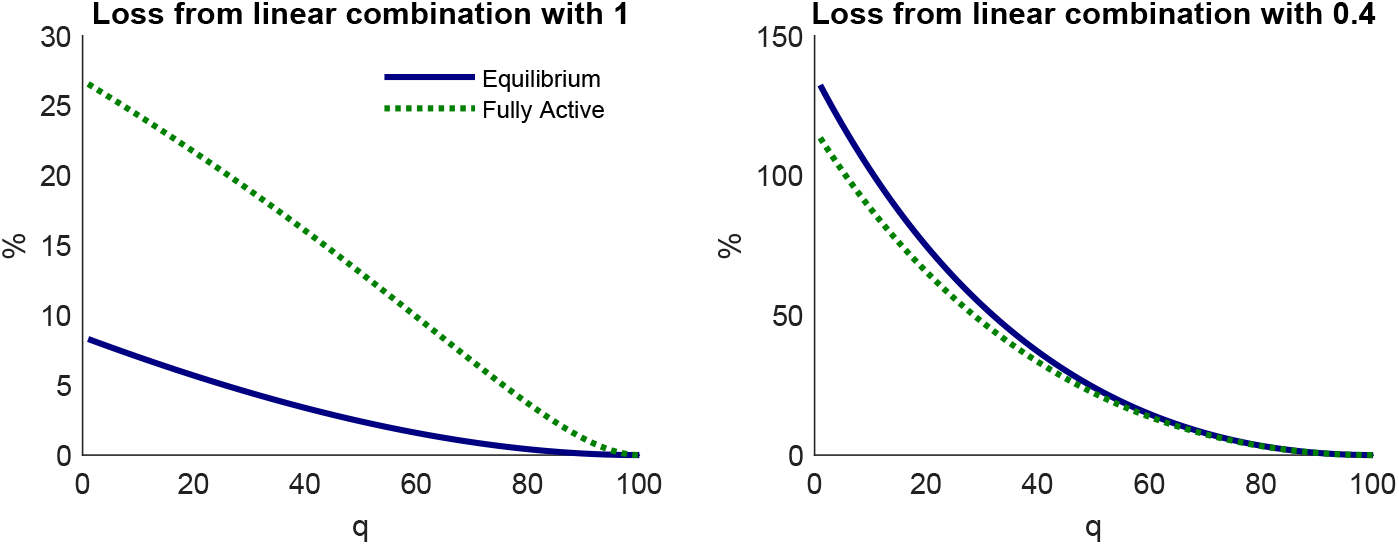
Global Optimality I.

In my final experiment, I randomly produce one million paths of 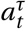 for the first 750 periods (the most relevant periods in my model) and use 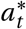 afterwards. Irrespective of *X*_*t*_ (equilibrium scenario on the left panel and fully active scenario on the right panel), Figure B3 suggests that a random path for 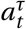 substantially (by more than 100%) reduces the value of the Hamiltonian.

**Figure B3:**
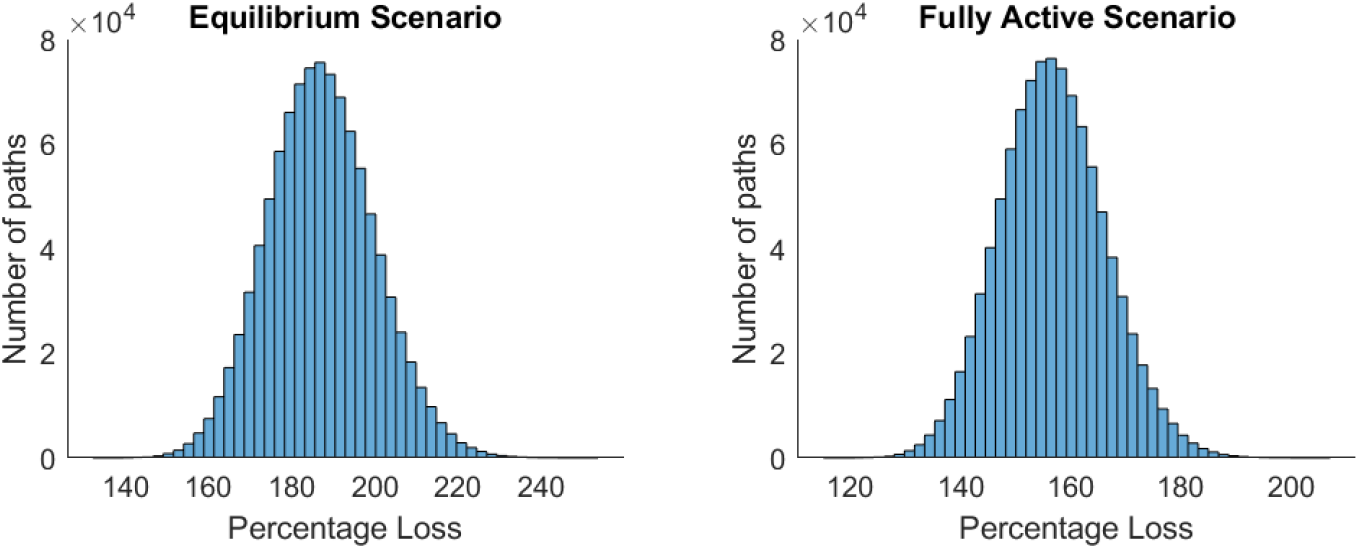
Global Optimality II.

All these experiments (and further experiments not reported) show that the optimal path generates a higher value of the Hamiltonian than multiple alternatives. As explained above, this does not prove that Eqs. (12-16) are sufficient for optimality. But these experiments do suggest that they are.

Finally, even though these results refer only to tested agents, similar conclusions apply to untested agents and the maximization of the Hamiltonian in Eq. (A2).

## C Single & Imperfect Testing - Further Results

### C.1 Cumulative Infections

**Figure B1:**
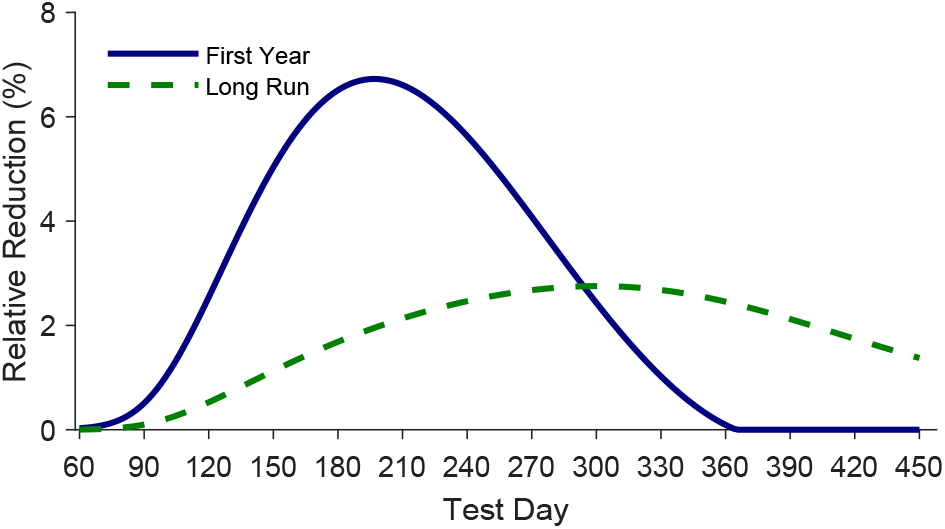
The Effect of the Test Day on Cumulative Infected. *Note:* This figure shows how the test day affects the reduction in cumulative infected caused by antibody testing in the first 12 months of the pandemic and in the long run relative to the scenario without tests (*ω* = 0). This figure assumes perfect testing.

### C.2 Welfare

**Table B1:** Benefits of Single & Imperfect Testing - Susceptible Agents’ Welfare.

| Test Day | Negative (%) |  |  |  | Positive (%) |  |  |  |
| --- | --- | --- | --- | --- | --- | --- | --- | --- |
|  | 60 | 150 | 300 | 450 | 60 | 150 | 300 | 450 |
| $p = q = 0$ | 0.01 | 3.05 | 16.67 | 20.55 | — | — | — | — |
| $p = q = 0.02$ | -0.00 | 2.59 | 15.61 | 19.33 | -0.01 | -5.55 | 9.94 | 16.76 |
| $p = q = 0.10$ | -0.00 | 1.46 | 11.61 | 14.55 | -0.00 | -2.33 | 6.71 | 12.19 |
| $p = 0.02; q = 0$ | -0.00 | 2.65 | 16.02 | 19.93 | -0.01 | -5.51 | 10.41 | 17.40 |
| $p = 0.10; q = 0$ | -0.00 | 1.72 | 13.71 | 17.63 | -0.00 | -2.37 | 8.90 | 15.33 |
| $p = 0; q = 0.02$ | 0.01 | 2.99 | 16.26 | 19.95 | — | — | — | — |
| $p = 0; q = 0.10$ | 0.01 | 2.73 | 14.68 | 17.66 | — | — | — | — |

*Note:* This table reports the percentage change in the shadow value of susceptible agents, $V_{s,T}^T$ , caused by widespread antibody testing on different test days, $T$ , relative to a benchmark without tests. On the left (right) panel, it reports the percentage change in the shadow value if the test is negative (positive). The four columns in each panel show how the results under different testing dates (from day 60 to day 450). Each line assumes a different combination of false positive, $p$ , and false negative, $q$ , rates.

**Table B2:** Benefits of Single & Imperfect Testing - Asymptomatic Agents’ Welfare.

| Test Day | Negative (%) |  |  |  | Positive (%) |  |  |  |
| --- | --- | --- | --- | --- | --- | --- | --- | --- |
|  | 60 | 150 | 300 | 450 | 60 | 150 | 300 | 450 |
| $p = q = 0$ | -0.00 | -0.48 | -0.40 | -0.10 | – | – | – | – |
| $p = q = 0.02$ | -0.00 | -0.53 | -0.41 | -0.10 | 0.17 | 2.65 | 0.46 | 0.05 |
| $p = q = 0.10$ | -0.00 | -0.59 | -0.40 | -0.09 | 0.03 | 2.08 | 0.44 | 0.05 |
| $p = 0.02; q = 0$ | -0.00 | -0.54 | -0.42 | -0.10 | 0.17 | 2.65 | 0.46 | 0.05 |
| $p = 0.10; q = 0$ | -0.00 | -0.67 | -0.48 | -0.11 | 0.03 | 2.16 | 0.45 | 0.05 |
| $p = 0; q = 0.02$ | -0.00 | -0.47 | -0.39 | -0.10 | – | – | – | – |
| $p = 0; q = 0.10$ | -0.00 | -0.42 | -0.34 | -0.08 | – | – | – | – |

*Note:* This table reports the percentage change in the shadow value of asymptomatic agents, $V_{e,T}^T$ , caused by widespread antibody testing on different test days, $T$ , relative to a benchmark without tests. See the note to Table B1 for more details.

**Table B3:** Benefits of Single & Imperfect Testing - Recovered Agents’ Welfare.

| Test Day | Negative (%) |  |  |  | Positive (%) |  |  |  |
| --- | --- | --- | --- | --- | --- | --- | --- | --- |
|  | 60 | 150 | 300 | 450 | 60 | 150 | 300 | 450 |
| $p = q = 0$ | – | – | – | – | 100.00 | 100.00 | 100.00 | 100.00 |
| $p = q = 0.02$ | -0.11 | -19.46 | -86.11 | -176.65 | 5.14 | 98.32 | 99.82 | 99.88 |
| $p = q = 0.10$ | -0.10 | -22.02 | -84.96 | -155.92 | 0.86 | 77.32 | 95.49 | 96.88 |
| $p = 0.02; q = 0$ | – | – | – | – | 5.24 | 98.39 | 99.83 | 99.89 |
| $p = 0.10; q = 0$ | – | – | – | – | 0.96 | 80.17 | 96.43 | 97.57 |
| $p = 0; q = 0.02$ | -0.06 | -17.30 | -82.73 | -173.27 | 100.00 | 100.00 | 100.00 | 100.00 |
| $p = 0; q = 0.10$ | -0.06 | -15.70 | -72.18 | -145.33 | 100.00 | 100.00 | 100.00 | 100.00 |

*Note:* This table reports the percentage change in the shadow value of recovered undocumented agents, $V_{f,T}^T$ , caused by widespread antibody testing on different test days, $T$ , relative to a benchmark without tests. See the note to Table B1 for more details.

## Footnotes

1 I contrasted the estimated number of immune agents and the number of individuals diagnosed. For the antibody surveys, see the ONS COVID-19 Infection Survey for the UK; for France and Spain, see Salje et al. (2020) and Pollán et al. (2020).

2 The human body can develop immunological memory (become immune) by developing special B and T cells, which defend the body against pathogens that it has previously encountered (Punt et al., 2018). Antibodies are produced by B-cells and are detected by antibody (serological) tests; but antibody tests do not detect T cells and it is theoretically possible that an individual is immune to the virus causing COVID-19 without developing antibodies thanks to T cells. As there is not yet, best to my knowledge, concrete evidence of that in the context of COVID-19, I simplify my analysis and abstract from the possibility that immune individuals do not develop antibodies.

3 To focus on how economic decisions under uncertainty affect the propagation of the COVID-19 pandemic, I abstract from more complex economic setups. For the latter, see, e.g., Bodenstein, Corsetti and Guerrieri (2020), Eichenbaum, Rebelo and Trabandt (2020*a*), and Krueger, Uhlig and Xie (2020), who include endogenous labor supply and differentiated goods implying different social-contact levels. Due to the pandemic, then, susceptible agents refrain from supplying labor and buying goods that imply much social contact, which in my model, are mechanisms captured in the agents’ choice of social activity.

4 Eichenbaum, Rebelo and Trabandt (2020*b*) differs, to some extent, from the others because they simultaneously assess the roles of viral and antibody testing. Yet, their approach does not allow to single out the role of antibody testing that I emphasize in this paper.

5 There is recent evidence suggesting that immunity against the virus causing COVID-19 is temporary (Seow et al., 2020). Despite that, I assume permanent immunity for three reasons. One is tractability: the time elapsed since agents learn that they lost immunity matters for their decision making; thus, there is growing heterogeneity in the model, which makes intractable to solve it. Most importantly, in early simulations of the COVID-19 pandemic, a vaccine was expected to emerge within 18 months (Ferguson et al., 2020) and have, in fact, been approved in about 10 months in December 2020. This is clearly below the 2-year average immunity duration against other coronaviruses (Huang et al., 2020), suggesting that vaccines are available when most recovered agents are still immune. Furthermore, Malkov (2020) and çenesiz and GuimarÃes (2020) using an SIRS (Susceptible-Infected-Recovered-Susceptible) model show, respectively, that waning immunity barely affects the epidemiological dynamics and optimal decentralized decision-making for about 18 months past the beginning of the epidemic. As I focus on the gains from antibody testing in the first 12 months, immunity duration appears almost irrelevant for my results.

6 Home antibody tests are already available for adults working on adult social care in England or domiciliary care in Wales (https://www.gov.uk/government/publications/coronavirus-covid-19-antibody-tests visited on 30 September 2020).

7 The way I model antibody testing is naturally a simplification but necessary for tractability as agents’ optimal decisions depend on their testing history. Eichenbaum, Rebelo and Trabandt (2020*b*) find a similar problem and make similar simplifying assumptions.

8 Papers assessing the implications of viral testing also abstract from imperfect testing (Berger, Herkenhoff and Mongey, 2020; Brotherhood et al., 2020; Eichenbaum, Rebelo and Trabandt, 2020*b*; Piguillem and Shi, 2020).

9 In reality, antibody surveys (similar to small *ω*) critically inform agents and governments about the extent of the pandemic. In the model, however, I abstract from these benefits of antibody tests to focus solely on the benefits that they generate by revealing whether agents are immune or not.

10 Brotherhood et al. (2020) and Piguillem and Shi (2020) make a similar assumption.

11 This cost of dying includes the foregone utility not associated with social activity (e.g., the utility associated with consumption and the utility from other activities that do not require social interactions). In a nutshell, the cost of dying is the value of life for the agent.

12 In Appendix B, I present numerical results suggesting that Eqs. (12-16) are also sufficient for optimality. See also Gersovitz and Hammer (2003) for a discussion of sufficiency conditions in economic models of epidemics and Goenka, Liu and Nguyen (2014, 2020) for formal proofs of sufficiency in the context of economic models of epidemics.

13 This evidence is reported in https://www.ft.com/content/033745f3-2d78-4869-9690-ea46fcc9cb3d, which I consulted on 17 June 2020.

14 There are some opposing indirect effects as well but they are of second-order importance. For example, tested agents restrain their social activity by less when *ω* is high as *X*_*t*_ is lower. Yet, these general-equilibrium effects do not change the qualitative results.

15 As immunity is assumed to be permanent, the pandemic asymptotically disappears when enough agents are immune.

16 In both cases, the cumulative number of infections exceed the herd immunity threshold, 1 − 1/*R*0. In my model, herd immunity is achieved when 60.6% of the individuals have been infected. This number assumes that symptomatic agents are always quarantined, i.e., *R*0 = *β*/*γ*_*e*_ + *σA*_*i*_ *β*/*γ*_*i*_.

17 Essentially, the distinction between announced and unannounced dates only slightly affects social activity prior to the test. Thus, I prefer the case of an unannounced date, which implies that tested and untested agents behave in the same way until the date of the test.

18 Figure B1 in the Appendix complements Table 2 by showing how the test day affects cumulative infections in the first year and in the long run. For example, this figure shows that on Day 200, when there is high health state uncertainty as more than 22% of the population is recovered undocumented and less than 66% is susceptible, antibody testing reduces cumulative infections by almost 7% in the first year.

19 This can be seen in two ways. First, as the test occurs later, the probability of being recovered undocumented on the date of the test is higher, leading to a larger fall in activity. Second, as most of the pandemic has already run its course after 300 days, asymptotically, the probability of being recovered undocumented does not subsequently increase as much, leading to a more persistent reduction in activity.

20 https://www.fda.gov/medical-devices/coronavirus-disease-2019-covid-19-emergency-use-authorizations-medical-devices/eua-authorized-serology-test-performance consulted on 25 September 2020. They report the test performance in terms of sensitivity, 1 − *p*, and specificity, 1 − *q*, of about forty antibody tests, discriminating in some cases between IgM and IgG sensitivity and specificity.

21 In the early days of the pandemic, the prior probability of being recovered undocumented is low, which leads to more false positives and, thus, a positive test indicates a lower posterior probability of being recovered undocumented.

